# Assessment of knowledge, attitude, and practice of respondents’ towards Rabies and associated risk factors in Shone Town, Hadiya Zone, Southern Ethiopia

**DOI:** 10.1101/2023.09.06.23295175

**Authors:** Teketel Gizaw Beresa, Teshita Edaso Beriso

**Author notes:** Corresponding author:, (TEB).

## Abstract

A survey-based cross-sectional study was carried out in the community of Shone Town, Hadiya Zone, South Region of Ethiopia, from November 2022 to April 2023 to assess respondents’ knowledge, attitude, and practice regarding rabies and associated factors. Woreda was selected purposefully, while kebeles and study populations were selected by simple random sampling. A total of 416 respondents were randomly selected and interviewed using a semi-structured questionnaire. All respondents (100%) heard about rabies from different sources, with the majority of them hearing about it from informal sources (62%), followed by mixed (mass media and traditional ways). 31.7% of those surveyed were aware that a virus was the cause of rabies. The findings revealed that 51.9% of individuals were aware of saliva contact, 0.7% were aware of rabid animal bites, and 47.4% were aware of both modes of transmission. With regard to the 100% fatal nature of rabies once the clinical signs developed, 64.4% of participants knew and the rest, 35.6%, did not. In terms of washing dog bite wounds with soap and water, the majority (86.3%) of respondents were aware. Of all participants, 73.1% agreed that stray dogs are dangerous, and 78.4% agreed that rabies is a problem for the community. With regard to the idea of killing stray dogs for the purpose of rabies prevention, 51.4% of respondents agreed that it was an effective method. 72.6% of respondents had contact with pets; however, 32% washed their hands after touching the pet animals. More than half of respondents practiced killing to control stray dogs. 36.8% of interviewees were experienced in vaccinating their dogs. In comparing the associations of several demographic risk factors with the mode of transmission and the risk of not vaccinating dogs, it was discovered that sex, age, occupations, and family size were statistically significant with both the risk of not vaccinating dogs and the means of transmission (P<0.05). The study demonstrates that a lack of formal education programs in the communities, low levels of education, and the majority of respondents acquiring their knowledge from unofficial sources were important contributors to the low levels of awareness among communities. The community should have been made aware of rabies through regular education, which should have been planned by veterinarians and medical professionals.

**Authors summary:** In Ethiopia, rabies is a leading cause of death that can be prevented. At Shone Town, Southern Ethiopia, we assessed community knowledge, attitudes, and practices toward rabies and its associated risk factors. All respondents heard about rabies from different sources, with most of them learned about it from informal sources. The participants were aware that the main ways of transmission were animal bites and saliva contact. The majority of participants agreed that stray dogs are harmful to the community and hazardous. More than half of those surveyed admitted to killing stray dogs. Most participants did not provide proper first aid after a suspected rabies bite, and contrary to accepted guidelines, the majority of animals were put to death rather than quarantined after a bite occurrence. The study shows that low levels of formal education programs in the communities and the fact that most respondents got their information from unreliable sources were major factors in the low levels of awareness among communities.

## 1. Introduction

Rabies is one of the neglected tropical diseases (NTDs), accounting for over 80% of human cases and primarily affecting poor and vulnerable communities [1]. *Lyssa virus* of the *Rhabdoviridae* family, which causes rabies, is a fatal infection that is endemic in developing countries, with Asia and Africa causing over 95% of human deaths [2]. Rabies infects all warm-blooded animals, and domestic dogs are responsible for up to 99% of rabies virus transmission to humans in endemic areas. Rabies is a virus that is carried by animals, primarily carnivores such as *canids* [3].

The most common means for rabies transmission are any types of bites, scratches, or other incidents in which saliva, cerebral or spinal fluid, tears, or nervous tissues from a suspected or known rabid animal or person enter an open wound, are transplanted into, or come into contact with the mucous membrane of another animal or person [4]. Rabies virus infection most commonly occurs when a rabid animal bites an animal or a person [5]. The virus then invades the central nervous system, resulting in acute and fatal encephalomyelitis [6]. In addition to its animal and public health significance, rabies also has significant economic importance globally, with the economic burden of dog-mediated rabies estimated at US$ 8.6 billion per year [7].

Vaccination, wound care, and the injection of rabies immunoglobulin are examples of preventive methods. However, in many developing countries, where canine rabies causes the majority of human cases, deaths occur mostly due to a lack of access to affordable biological agents needed for effective post-exposure prophylaxis. A reduction in the number of human deaths due to rabies has to begin with the elimination of canine rabies. The feasibility of eliminating canine rabies in Africa is predicated on understanding and counteracting the many reasons that canine rabies control has failed in Africa. Domestic dog vaccination offers a cost-effective strategy for the prevention and elimination of human rabies mortality, and it is epidemiologically and practically feasible to eradicate canine rabies through mass vaccination of domestic dogs [8].

The WHO has established the “zero by 30” global strategy plan, which aims to eradicate endemic rabies globally by 2030 by preventing human deaths caused by dog-mediated rabies [9]. Even though widespread vaccination is the most well-known and effective strategy, there is still a lot that can be done to make it even more effective. Economic, cultural, social, educational, and technological issues must also be considered, particularly in Asia and Africa, where the rabies burden is significant, including Ethiopia [10].

Ethiopia ranks fourth on the globe and has the second-highest number of rabies mortality rates on the African continent, after Nigeria [11]. The disease has been ranked first among the top five zoonotic diseases (rabies, anthrax, brucellosis, leptospirosis, and echinococcosis) in Ethiopia by a panel of experts from human, animal, and environmental health [12, 13]. Due to a large dog population that is poorly managed, rabies has been recognized as the most common disease in Ethiopia for many centuries [14]. It is primarily a disease of dogs in the country because access to suspected domestic canids and pets is not controlled indoors or by immunization. In the past two decades, a high number of animal rabies cases have been confirmed in Addis Ababa, the capital city of Ethiopia, and the majority of rabies cases were confirmed in dogs [15].

Understanding communities’ perceptions of the cause, mode of transmission, symptoms, treatment, and possible intervention measures of rabies is an important step towards developing strategies aimed at controlling the disease, determining the level of implementation of planned activities in the future, and creating responsible pet ownership, routine veterinary care and vaccination, and professional continuing education [16]. Poor public awareness of rabies is considered one of the challenges to disease prevention and control in Ethiopia, including the study area. Therefore, the objectives of this study were:

❖ To assess the level of knowledge, attitude, and practices of shone town communities towards rabies in Hadiya Zone, South Ethiopia.
❖ To identify factors associated with community knowledge, attitude, and practice about rabies in the study area.

## 2. Materials and methods

### 2.1 Study area

The survey was conducted in Shone town, which is the administrative center of Misrak Badewacho Woreda. Misrak Badewacho Woreda is located in the Hadiya Zone of the Southern Nations, Nationalities, and People Regional State (SNNPRS). Because Kambata Tembaro zone exists between Misrak Badewacho and the other Woredas of Hadiya zone, Misrak Badewacho Woreda has a unique geographic feature in that it does not share a boundary with any other Woredas of Hadiya zone, with the exception of Mirab Badewacho Woreda. Geographically, Misrak Badewacho is located between 07° 03′ 20′′ N and 07° 16′ 08′′ N of latitudes and 037° 53′ 02′′ E and 038° 06′ 02′′ E of longitudes. It is bounded by Alaba zone in the North, Siraro Woreda of Oromiya region in the East, Kedida Gamela Woreda in Kambata Tembaro zone, Mirab Badewacho Woreda in the West, and Damot Gale and Damot Pulasa woreda of Wolaita zone in the South. It is located at a distance of 354 km south-west of Addis Ababa, the capital city, along the road from Addis Ababa through Shashemane to Wolaita Sodo. It is also found at a distance of 123 km and 97 km from Hawassa, the regional capital city, and Hosanna, the zone capital town, respectively [17].

### 2.2 Study population

The study populations were individuals who were residents of Shone town with different socio-demographic characteristics. Shone town has six kebeles and the study includes both sexes of individuals, respondents above the age of seventeen, and households that have been residents of the area for at least four months, with different occupations and different marital statuses. Besides, the target populations were interviewed with specific questions related to the assessment, knowledge, attitude, and practice of the community regarding rabies.

### 2.3 Study design

A cross-sectional questionnaire-based study design was conducted by collecting data through structured questionnaires.

### 2.4 Sample size determination and sampling technique

The Shone town was selected purposefully, while the kebeles and study populations were chosen by simple random sampling. The sample size was determined based on the formula developed by Yamane (1967), assuming a 95% confidence interval and a 0.05 sampling error. Population size was estimated at 5500; e = sampling error = 0.05.

n = N/1+ (N (e) 2) =5500/1+500(0.05*0.05) = 5500/1+5500*0.0025=373, n = number of people groups exposed to pet or other domestic animal bites. But to increase precision, 420 respondents were selected for the study.

#### 2.4.2 Sampling procedures

Face-to-face interviews are carried out using structured and pretested questionnaires. The questionnaire contains levels of KAP with respect to rabies management and control, household information, and pet care. Animal vaccination will be defined as having been immunized (oral or parenteral) one year before the survey. Prior to the pretesting and survey, the questionnaires had been translated into the local language (Hadyisa) and national language (Amharic) and then back-translated to English to ensure validity.

### 2.5 Method of data collection

A semi-structured questionnaire has been designed to collect information about the respondents’ knowledge of the rabies disease, its cause, means of transmission, treatment, and prevention practices. Included in the demographic variables were 39 questions (16 knowledge, 7 attitudes, and 10 practices) and 7 demographics asked of each participant regarding the cause, sources, and mode of transmission, attitude, practice, and prevention and control measures of rabies. Respondents above the age of seventeen and households that have been residents of the area for at least four months were included. The questionnaire was prepared in English and Amharic, then translated to the local language for appropriateness and ease of approaching the respondents. The study participants were asked for their consent orally and interviewed face-to-face in their homes by instilling a semi-structured (closed and open-ended) questionnaire through a handful of house-to-house surveys of residents in all six kebeles of Shone town.

### 2.6 Data management and analysis

The collected raw data were stored, coded in a personal computer’s Microsoft Excel spread sheet program, and analyzed using STATA statistical software version 14. Descriptive statistics such as frequencies, distributions, and percentages were used to summarize the data. Pearson’s chi-square test was used to detect the existence of an association between different risk factors and outcome variables. Besides, a p-value of less than 0.05 was considered statistically significant.

## 3. Result

### 3.1 Socio-demographic characteristics

A total of 420 respondents were interviewed for assessment of knowledge, attitude, and practice towards rabies and associated risk factors in Shone Town. Among the data collected, 4 respondents were found incomplete and excluded from the analysis, and then the remaining data was analyzed. Out of 416 respondents, 52.6.1% and 47.4% were males and females, respectively. Respondents with an age greater than 65 years were 4.3%, between 51 and 65 years were 16.3%, 31–50 years were 32.2%, and 18–30 years were 47.1% (Table 1).

**Table 1:** Socio-demographic characteristics of participants.

| Variables | Category's | Number of respondents | Frequency (%) | 95% CI |
| --- | --- | --- | --- | --- |
| Sex | Male | 219 | 52.6 | 47.81-57.42 |
|  | Female | 197 | 47.4 | 42.57-52.18 |
| Age (in years) | 18-30 | 196 | 47.1 | 42.34-51.94 |
|  | 31-51 | 134 | 32.2 | 27.87-36.87 |
|  | 51-65 | 68 | 16.3 | 13.08-20.23 |
|  | >65 | 18 | 4.3 | 2.73-6.77 |
| Marital Status | Single | 157 | 37.7 | 33.18-42.51 |
|  | Married | 251 | 60.3 | 55.53-64.94 |
|  | Divorced | 8 | 1.9 | 0.96-3.80 |
| Occupation | Un-employer's | 51 | 12.3 | 9.4 -15.79 |
|  | Self-employer's | 184 | 44.2 | 39.50-49.06 |
|  | Civil servant's | 53 | 12.7 | 9.85-16.31 |
|  | Student's | 70 | 16.8 | 13.51-20.75 |
|  | Farmer's | 58 | 13.9 | 10.92-17.63 |
| Educational status | No formal education | 37 | 8.9 | 6.50-12.05 |
|  | Primary education | 101 | 24.3 | 20.38-28.65 |
|  | higher education | 104 | 25.0 | 21.05-29.40 |
|  | Secondary education | 174 | 41.8 | 37.15-46.64 |
| Family member | 1-3 | 113 | 27.2 | 23.08-31.66 |
|  | 4-6 | 95 | 22.8 | 19.03-27.13 |
|  | >6 | 208 | 50.0 | 45.19-54.80 |
| Live with pets animals | Live without pets | 130 | 31.3 | 27.34-34.71 |
|  | Live with a pet | 286 | 68.8 | 55.03-71.23 |

### 3.2 Knowledge of interviewees toward rabies

All interviewed respondents were heard about rabies from different sources, that majority from non-mass media 258(62%), followed by both mass media and traditional ways and respondent had familiar with local name ‘Wisha gereechiso jabo’ or ‘Wisha machaso jabo’ by Hadiysa or **‘**Yebade wusha bashita’ in national language Amharic. 31.7% respondents knew that cause of rabies was virus and the rest 68.3 respondents knew starvation and others as causes. All respondents knew as all warm blooded animal included human being infected by rabies and dog is main source of rabies. Out of all, 51.9% participants knew bite of rabid animals, 0.7% saliva contact and 47.4% by saliva contact and bite of rabid animal as mode of transmission (Table 2).

**Table 2:**
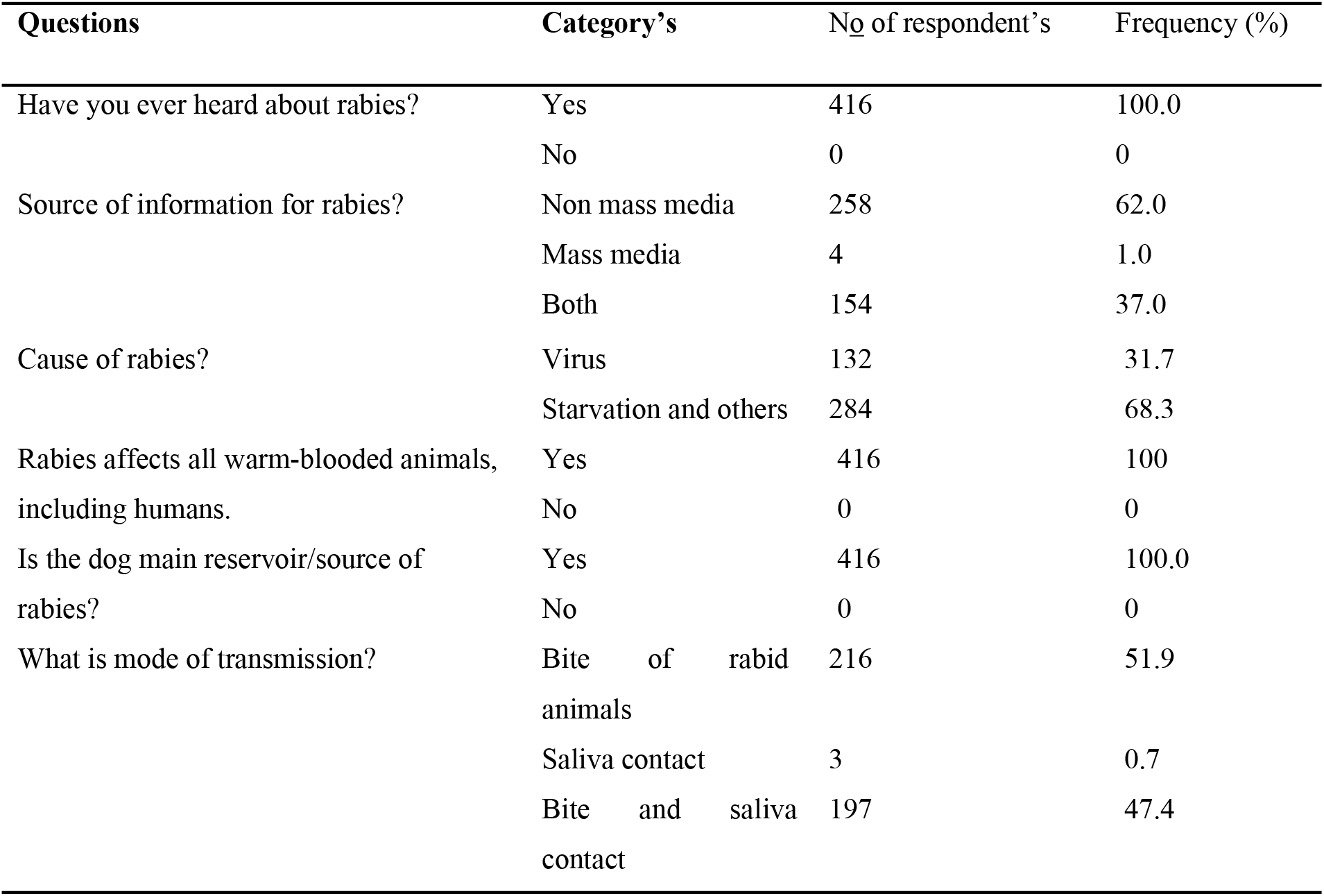
Assessment of knowledge of the respondents on rabies.

| Questions | Category's | No of respondent's | Frequency (%) |
| --- | --- | --- | --- |
| Have you ever heard about rabies? | Yes | 416 | 100.0 |
|  | No | 0 | 0 |
| Source of information for rabies? | Non mass media | 258 | 62.0 |
|  | Mass media | 4 | 1.0 |
|  | Both | 154 | 37.0 |
| Cause of rabies? | Virus | 132 | 31.7 |
|  | Starvation and others | 284 | 68.3 |
| Rabies affects all warm-blooded animals, including humans. | Yes | 416 | 100 |
|  | No | 0 | 0 |
| Is the dog main reservoir/source of rabies? | Yes | 416 | 100.0 |
|  | No | 0 | 0 |
| What is mode of transmission? | Bite of rabid animals | 216 | 51.9 |
|  | Saliva contact | 3 | 0.7 |
|  | Bite and saliva contact | 197 | 47.4 |

Majorities (80.3%) of interviewees knew about clinical signs, and some respondents (19.7%) didn’t. Out of all respondents, 64.4% were aware of the lethal behavior of rabies, and the rest, 35.6%, were not. The majority (64.4%) of participants knew about the almost 100% fatal nature of rabies once the clinical signs developed. Most respondents (86.3%) wash dog bite wounds with soap and water (Table 3).

**Table 3:** Perceptions of participants about further rabies disease.

| Questions | Category's | No of participants | Frequency (%) |
| --- | --- | --- | --- |
| Clinical signs of the rabies disease | Know | 334 | 80.3 |
|  | Don't know | 82 | 19.7 |
| Do you know rabies can cause death in humans, dogs, and other domestic animals? | Yes | 412 | 99.0 |
|  | No | 4 | 1.0 |
| Can rabies be prevented by vaccination? | Yes | 412 | 99.0 |
|  | I don't know | 4 | 1.0 |
| Vaccine effectiveness after a suspected animal bite? | I don't know | 40 | 9.6 |
|  | Immediately | 376 | 90.4 |
| Can rabies be treated with post-exposure prophylaxis? | Yes | 254 | 61.1 |
|  | I don't know | 162 | 38.9 |
| Do you know the 100% fatal nature of rabies once the clinical signs develop? | Yes | 268 | 64.4 |
|  | No | 148 | 35.6 |
| Do you know the risk of getting rabies if the dog is not vaccinated? | Yes | 369 | 88.7 |
|  | No | 47 | 11.3 |
| Is it good to wash dog bite wounds with soap and water? | Yes | 359 | 86.3 |
|  | No | 47 | 11.3 |
|  | I don't know | 10 | 2.4 |

### 3.3 Attitude of respondents towards rabies

From the total participants, 73.1% had a positive attitude towards stray dog danger, while 16.6% did not. 78.4% of respondents agreed that rabies is a problem for the community, and 91.1% did not agree that holy water can treat rabies disease. With regard to the idea of killing stray dogs for the purpose of rabies prevention, 51.4% of respondents agreed that it was an effective method. All (100%) interviewees agreed that educating society has a great impact on the prevention of rabies (Table 4).

**Table 4:** Attitude of respondents towards rabies.

| Opinion's | Category's | No of respondent's | Frequency (%) |
| --- | --- | --- | --- |
| Stray dogs are dangerous | Agree (+ve attitude) | 304 | 73.1 |
|  | Neutral (no opinions) | 69 | 16.6 |
|  | Disagree (-ve attitude) | 43 | 10.3 |
| Is rabies an issue in your Keble for your community? | Agree (+ve attitude) | 326 | 78.4 |
|  | Neutral (no opinions ) | 49 | 11.8 |
|  | Dis agree (-ve attitude) | 41 | 9.9 |
| Rabies can be effectively prevented by euthanizing (killing) stray dogs. | Agree (+ve attitude) | 214 | 51.4 |
|  | Neutral (no opinions) | 114 | 27.4 |
|  | Dis agree (-ve attitude) | 88 | 21.2 |
| Do you believe holy water can treat rabies disease? | Agree(-ve attitude) | 37 | 8.9 |
|  | Disagree (+ve attitude) | 379 | 91.1 |
| Celebrating World Rabies Day every year on September 28 has its own role in reducing reduction cause of rabies disease in animals and people. | Agree (+ve attitude) | 311 | 74.8 |
|  | Neutral (no opinions ) | 102 | 24.5 |
|  | Dis agree (-ve attitude) | 3 | 7 |
| Rabies can be prevented by educating people about the disease. | Agree (+ve attitude) | 416 | 100.0 |
|  | Disagree (-ve attitude) | 0 | 0 |
| Willing to register pets? | Agree (+ve attitude) | 308 | 74.0 |
|  | Neutral (no opinions) | 108 | 26.0 |

### 3.4 Practice of respondents towards rabies

From the total respondents, 72.6% had contact with pets. However, from that, 32% had the habit of washing their hands after touching pet animals, and 21% had a history of being bitten by dogs. 100% of respondents had the perception that they should go to the hospital in case they were bitten by a pet animal. More than half of respondents practiced killing to control stray dogs. With regard to vaccination, 36.8% of them practiced vaccinating their dogs and had vaccination certificates (Table 5).

**Table 5:** Practices of respondents toward rabies disease.

| Questions | Category's | No of respondent's | Frequency (%) |
| --- | --- | --- | --- |
| Do any of your family members touch your dog or cat? | Yes | 302 | 72.6 |
|  | No | 114 | 27.4 |
| Do you wash your hands after touching the dog or cat? | Yes | 133 | 32 |
|  | No | 283 | 68 |
| Have you ever been bitten by a dog? | Yes | 73 | 21.3 |
|  | No | 343 | 78.7 |
| What measures do you take to control stray dogs? | Killing | 220 | 52.9 |
|  | aware of the owner | 196 | 49.1 |
| Would you inform the authorities if you bitten by a dog? | Yes | 355 | 85.3 |
|  | No | 61 | 14.7 |
| Did you vaccinate your dog? | Yes | 153 | 36.8 |
|  | No | 263 | 63.2 |
| Can you show the dog's vaccination certificate? | Yes | 153 | 36.8 |
|  | No | 263 | 63.2 |
| Where do you care for your dog? | Free to move any | 75 | 18 |
|  | kept indoor | 211 | 50.3 |
|  | Other specify | 130 | 31.3 |
| Does dog registration and licensing help in the control of rabies? | Yes | 364 | 87.5 |
|  | No | 52 | 12.5 |

The perceptions of respondents about modes of transmission among different risk factors were assessed, and it was found that 34.7% and 46.19% of female and male interviewees knew that rabies was transmitted by bite of a rabid animal and saliva contact, respectively. Sex, age, occupations, and family size were statistically significant (p<0.05), when compared with mode of transmission (Table 6).

**Table 6:** Association of demographic characteristics with mode of transmission.

| Variables | Category | Mode of transmission | | | $\chi^2$ | P - Value |
| --- | --- | --- | --- | --- | --- | --- |
|  |  | Bite of rabid animal | Saliva contact | Bite and saliva contact |  |  |
| Sex | Male | 60.27 | 5.02 | 34.70 | 14.55 | 0.001 |
|  | Female | 42.64 | 11.17 | 46.19 |  |  |
| Age | 18-30 | 51.53 | 8.67 | 39.80 | 100.44 | 0.001 |
|  | 31-50 | 35.82 | 11.94 | 52.24 |  |  |
|  | 51-65 | 1.47 | 0.00 | 98.53 |  |  |
|  | >65 | 0.00 | 0.00 | 100.00 |  |  |
| Marital status: | Single | 53.50 | 10.19 | 36.31 | 4.22 | 0.376 |
|  | Married | 51.39 | 6.77 | 41.83 |  |  |
|  | Divorced and other | 51.92 | 7.93 | 40.14 |  |  |
| Occupation | Unemployed | 72.55 | 23.53 | 3.92 | 155.50 | 0.000 |
|  | Self-employed | 49.46 | 8.70 | 41.85 |  |  |
|  | Civil servant | 9.43 | 0.00 | 90.57 |  |  |
|  | Student | 35.71 | 7.14 | 57.14 |  |  |
|  | Farmer | 100.00 | 0.00 | 0.00 |  |  |
| Family members | >6 | 62.02 | 0.48 | 37.50 | 120.56 | 0.000 |
|  | 4-6 | 44.21 | 32.63 | 23.16 |  |  |
|  | <4 | 39.82 | 0.88 | 59.29 |  |  |

The knowledge of respondents about the risk of not vaccinating dogs was compared with different demographic characteristics. Females (93.91%), age groups between 35 and 50 years (100%), and civil servants (100%) had good perceptions of the risk of not vaccinating dogs. From demographic risk factors, sex, age, occupations, and family size were found statistically significant (P<0.05) (Table 7).

**Table 7:**
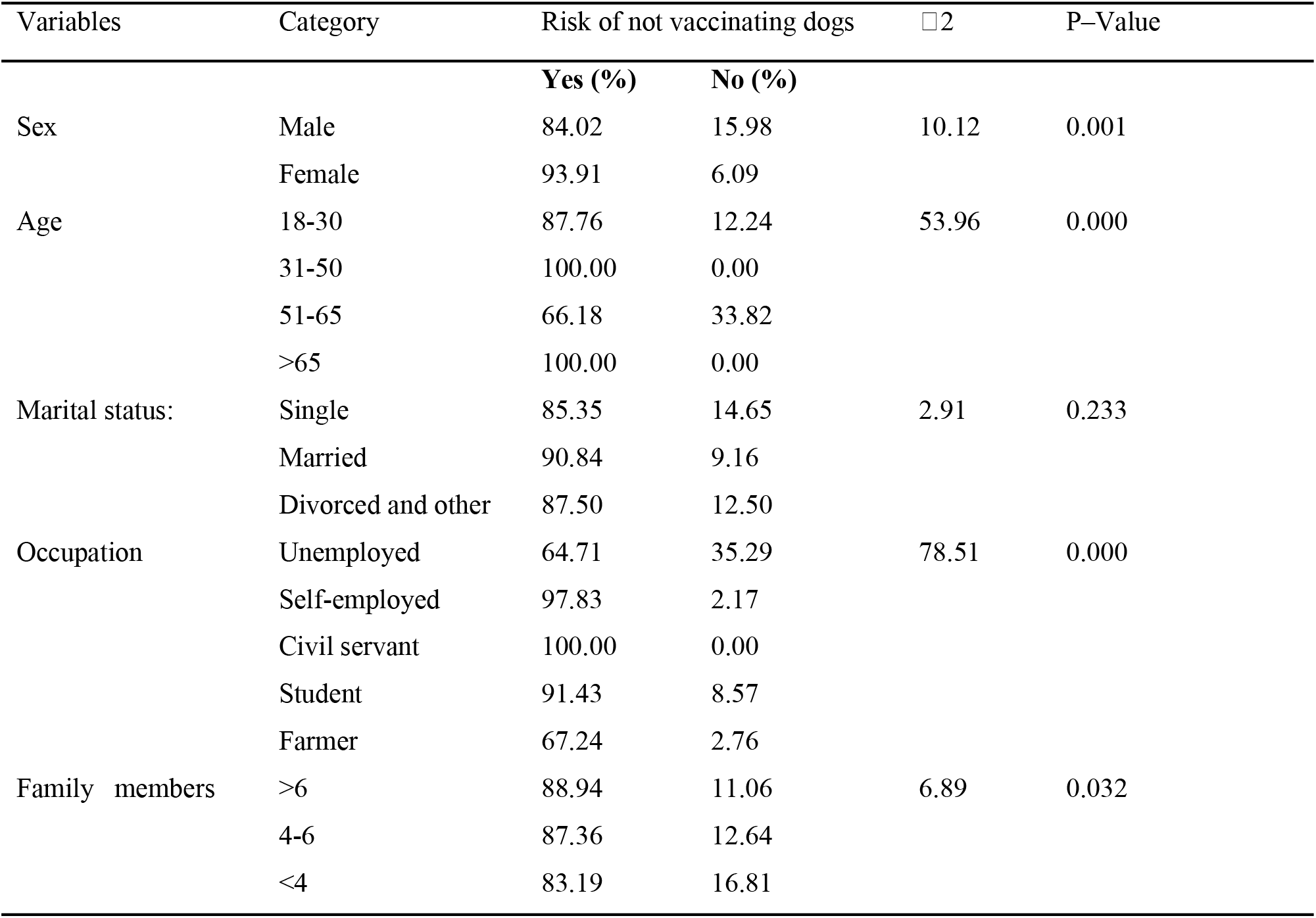
Association of demographic characteristics with the risk of not vaccinating dogs.

## 4. Discussions

Rabies is a severe public health concern because of the disease’s rapid mortality rate, particularly in tropical nations like Ethiopia. Few awareness-raising and public education initiatives are conducted globally because rabies is regarded as a neglected tropical disease. According to preliminary research, the highest dog population is found in Ethiopia, where there is a higher risk of dog bites and rabies. The present study revealed that all interviewed respondents had heard about rabies from different sources 416 (100%). The result of this study was consistent with the report that most respondents had heard [18]. Contrary to the present study in Mersa town, in which the majority of respondents (83.33%) had not heard about rabies [14]. The current study revealed that 100% of interviewees had heard about rabies. Regarding the source of information, 62.0% of the respondents heard about rabies from non-mass media (from their family and neighbors), 1% from formal media (media, animal health workers), and 37% heard from both sources. This discrepancy is most likely explained by the absence of rabies-related media coverage and community education in the research area.

In the current study, 31.7% of the respondents knew that a virus is the cause of rabies. This was higher than the finding of [19], who reported that 18% of the respondents knew that virus was the causal agent of rabies, and 25.83% of respondents knew virus was the cause of rabies [14]. Regarding the causative agent of rabies, the majority of respondents were found to have misperceptions, which were starvation and thirst. This result is consistent with a study in Munesa District, Arsi Zone, Southeastern Ethiopia [20] that reported misperceptions of starvation and thirst as causes of rabies. Similar misperceptions were reported by several scholars from different areas of Ethiopia [21-23]. However, this variation might be associated with a difference in community awareness between different study areas. The possible reason for this could be due to the lack of information gained by the media about rabies causes.

This finding revealed that all respondents knew that all warm-blooded animals, including humans, are infected by rabies, and dogs are the main source of rabies. The result of this study was consistent with the report from Mozambique that the majority (96.5%) of participants were aware that dogs are the main source of rabies and that dog could be affected by the disease (98.2%) [24]. Out of all, 51.9% of participants knew bites of rabid animals, 0.7% saliva contact, and 47.4% saliva contact and bites of rabid animals as modes of transmission. Most of the respondents tried to identify signs and symptoms of the rabies disease in both animals and humans.

In the current study, 13.7% of respondents did not know about post-exposure treatment, including washing the bite site wound with water and soap. Contrary to the present study, about 44% of respondents did not know that post-exposure treatment starts with nonspecific treatment such as washing the site of the bite with water and disinfectant [25]. The percentage of respondents who were unaware was consistent when compared to other reports, such as 18% in Cameroon reported by Barbosa and 30.7% in rural household heads of Gondar Zuria District, Ethiopia, reported by Digafe [26, 27]. However, this percentage of respondents who were unaware is high when compared to Bangladesh, at 2% [28]. In the current study, 88.7% knew the risk of getting rabies if the dog was not vaccinated. In contrast, 57.5% of the study participants did not know that keeping dogs that are not vaccinated against rabies is dangerous [25]. This may be due to the difference in frequency and method of information dissemination from community to community.

Among the study participants, 36.8% had vaccinated their dogs. In contrast, a high percent of study participants who vaccinate their dogs was reported from different studies conducted: 79% in Mekelle City, Ethiopia [29], and 71.1% in Adigrat Town, Tigray Regional State [30]. 85.3% of dog owners vaccinated their dogs in Chiro town, West Hararghe, Oromia region, Ethiopia [31]. The result of the recent study was greater than the study conducted in Jima [27]. The variation may be due to the availability of animal vaccines or conditions in the information sharing in the study area. In the present study, 18% of respondents managed their dogs by keeping them free. Lower than that, 60.83% of respondents managed their dogs by letting them run free [14]. This difference may be due to information about the importance of a control dog in reducing the risk of rabies.

## 5. Conclusions and recommendations

The current study revealed that rabies is a significant disease in both humans and animals in the study area. The level of knowledge about the fatal nature and clinical signs of rabies in animals and humans is good. However, several knowledge gaps on the cause and mode of transmission of rabies were found. The participants had a positive attitude towards stray dog danger, as rabies is a problem for the community, and holy water cannot treat rabies disease. With regard to the idea of killing stray dogs for the purpose of rabies prevention, respondents agreed that this was an effective method. The majority of interviewees agreed that educating society has a great impact on preventing rabies. However, they lack the practice of washing their hands after touching the dog(s) or cat(s). Some respondents allow their dog to roam freely, putting their family and other domestic animals at risk of rabies. The incidence of rabies increased due to factors like a lack of community awareness, vaccination coverage, and regulation for stray dog control, as well as coordination mainly between the health and veterinary sectors as well as the community in the study area. Moreover, the majority of respondents were aware of any first line of action after an animal bite, including immediate visits to health facilities. The study illustrates that low levels of education, most respondents getting information from informal sources (family, friends), and a lack of formal education programs in the communities were major factors contributing to the poor awareness among communities. Based on the above conclusive points, the following recommendations were forwarded:

❖ Veterinarians and health professionals should have created a strategy for periodic education to raise community awareness about rabies.
❖ Every individual should have immediately washed their dog bite wounds with soap and water and gotten to the hospital.
❖ Every community pet’s owner should control their pet indoors and vaccinate.

## Data Availability

The analyzed data during this study will be provided on request from the corresponding author.

## Author Contributions

TEB and TG: conception, study design, execution, acquisition of data, analysis, and interpretation, drafting, revising, or critically reviewing the article; Both authors took part in drafting, revising, or critically reviewing the article; gave final approval of the version to be published; have agreed on the journal to which the article has been submitted; and agree to be accountable for all aspects of the work.

## Data Sharing Statement

The analyzed data during this study will be provided on request from the corresponding author.

## Ethical Clearance

The purpose of the survey was explained to participants, and the best practices for veterinary care were followed. The verbally informed consent procedure described in the consent and the study protocols with reference number WSU 41/22/2241 were both approved by the Wolaita Sodo University of Research Ethics and Review Committee.

## Consent for publication

Both authors read the final text and agreed to submit the manuscript in its present form.

